# A systematic review of the data, methods and environmental covariates used to map *Aedes*-borne arbovirus transmission risk

**DOI:** 10.1101/2023.04.19.23288781

**Authors:** Ah-Young Lim, Yalda Jafari, Jamie M. Caldwell, Hannah E. Clapham, Katy A. M. Gaythorpe, Laith Hussain-Alkhateeb, Michael A. Johansson, Moritz U. G. Kraemer, Richard J. Maude, Clare P. McCormack, Jane P. Messina, Erin A. Mordecai, Ingrid B. Rabe, Robert C. Reiner, Sadie J. Ryan, Henrik Salje, Jan C. Semenza, Diana P. Rojas, Oliver J. Brady

## Abstract

**Background:** *Aedes (Stegomyia)*-borne diseases are an expanding global threat, but gaps in surveillance make comprehensive and comparable risk assessments challenging. Geostatistical models combine data from multiple locations and use links with environmental and socioeconomic factors to make predictive risk maps. Here we systematically review past approaches to map risk for different *Aedes*-borne arboviruses from local to global scales, identifying differences and similarities in the data types, covariates, and modelling approaches used.

**Methods:** We searched on-line databases for predictive risk mapping studies for dengue, Zika, chikungunya, and yellow fever with no geographical or date restrictions. We included studies that needed to parameterise or fit their model to real-world epidemiological data and make predictions to new spatial locations of some measure of population-level risk of viral transmission (e.g. incidence, occurrence, suitability, etc).

**Results:** We found a growing number of arbovirus risk mapping studies across all endemic regions and arboviral diseases, with a total of 183 papers published 2002-2022 with the largest increases shortly following major epidemics. Three dominant use cases emerged: i) global maps to identify limits of transmission, estimate burden and assess impacts of future global change, ii) regional models used to predict the spread of major epidemics between countries and iii) national and sub-national models that use local datasets to better understand transmission dynamics to improve outbreak detection and response. Temperature and rainfall were the most popular choice of covariates (included in 50% and 40% of studies respectively) but variables such as human mobility are increasingly being included. Surprisingly, few studies (22%, 33/148) robustly tested combinations of covariates from different domains (e.g. climatic, sociodemographic, ecological, etc) and only 48% of studies assessed predictive performance via out-of-sample validation procedures.

*Conclusions:* Here we show that approaches to map risk for different arboviruses have diversified in response to changing use cases, epidemiology and data availability. We outline specific recommendations for future studies regarding aims and data choice, covariate selection, model formulation and evaluation.

**Author Summary:** *Aedes*-borne arboviruses such as dengue, Zika, chikungunya, and yellow fever pose a growing global threat. It is crucial to map their risk to target interventions and control their spread. A review of 183 studies found that risk mapping methods have evolved over time to respond to changing epidemiology and data availability. Initially, mapping risk involved using data from multiple areas and satellite imagery to develop models predicting transmission risk on a global or continental scale. Following Zika and chikungunya epidemics, mechanistic models based on national-level incidence data have been utilised to track the spread of epidemics across countries. The use of case-based surveillance systems has enabled more precise and detailed predictions at sub-national levels. Of the studies reviewed, half included temperature and rainfall as covariates, and human mobility was increasingly accounted for in arbovirus risk mapping. However, only 33 of the 148 studies robustly selected the variables included in their predictions, and only half of the studies assessed their accuracy against new data. The review suggests that future risk mapping studies should consider the purpose of the map, data quality, and methodological innovations to improve accuracy of risk maps to ensure they are useful for informing control of *Aedes*-borne arboviruses.

## Background

Arboviruses, commonly referred to as arthropod-borne viruses, are a wide range of viral pathogens transmitted through the bite of arthropods such as mosquitoes and ticks. The term arbovirus does not refer to a distinct taxonomic group, but the viruses have similar transmission mechanisms, which makes information gained from one virus potentially useful in understanding and preventing the spread of other viruses [1]. In this paper, we focus on *Aedes (Stegomyia)*-borne arboviruses, including dengue, Zika, chikungunya, and yellow fever, which are of particular concern due to their high disease burden and life-threatening health consequences [2]. The geographical spread and burden of this group of arboviruses have been rapidly increasing in recent years. It has been estimated that 100-400 million dengue infections occur each year worldwide, mainly in South America and South-East Asia (SE Asia), with the disease threatening to spread to new regions including Europe [3–5]. Zika and chikungunya viruses were first identified in Africa and Asia, but emerged and rapidly spread throughout the Americas between 2013 and 2015, likely due to a combination of suitable climatic factors, increasing international air travel and possible immunological drivers [6,7]. The Zika outbreak received global attention due to its link to congenital and neurological complications, resulting in the declaration of a Public Health Emergency of International Concern by the World Health Organization (WHO) in 2016 [7]. Chikungunya is frequently accompanied by joint pain and rheumatic manifestations that can persist for a long time and have a significant impact on the quality of life of affected individuals [4]. Yellow fever is endemic in tropical and subtropical countries of South America and Africa, with an estimated number of 109,000 severe infections and 51,000 deaths in 2018 [8]. Among the *Aedes*-borne arboviruses, yellow fever is the only one that has a safe and effective vaccine available for humans. A sylvatic cycle between non-human primate reservoirs and mosquitoes is the most common source of yellow fever virus infection; however, humans can also become infected through the urban cycle, which can potentially lead to large outbreaks, as recently seen in Angola, Nigeria and the Democratic Republic of the Congo [8,9]. As these *Aedes-*borne arboviruses share a common mechanism of transmission, the WHO launched the Global Arbovirus Initiative in 2022, which includes the aim of developing a comprehensive risk monitoring and early detection tool that will allow countries to assess global risk of different *Aedes*-borne viruses, strengthen vector control, and develop global systems and strategies to monitor and reduce the risk in the local, regional, and national levels. This initiative identified reviewing the drivers of spatial arbovirus risk at global and regional levels as a key priority.

Surveillance of arboviral diseases varies among countries, by clinical manifestations, and over time, but three main data types are used most commonly for risk mapping: disease occurrence, case incidence, and seroprevalence data. Occurrence data represent a specific location where one or more cases of a disease has occurred [10] (e.g. an outbreak report) and is often available even in otherwise data-sparse regions, but conveys limited information about the magnitude of risk. Case incidence, as measured by traditional, largely passive disease surveillance systems, provides more information on magnitude due to being denominator-based (e.g. cases per 1,000 residents), but often underestimates the incidence of infection and is often not directly comparable between countries due to differing case definitions, health seeking patterns, health care and laboratory capacity, immunological landscape and surveillance systems. Age-specific community-representative seroprevalence survey data, when combined with models, can be used to estimate force of infection. This provides a less biased measure of long-term transmission risk, but is the least abundant data type and is subject to the limitations of serology in the context of cross-reactive flavivirus infections [11].

The geographic distribution and intensity of *Aedes*-borne arbovirus transmission have been attributed to a combination of pathogen, environmental, demographic and socioeconomic factors such as climate change, urbanisation and local and international travel. Temperature, in particular, is a frequently cited determinant of arbovirus transmission, as temperature drives all important metabolic traits for vector mosquitoes to transmit the virus to humans [12]. Rapid unplanned urbanisation increases human population density, can create urban heat islands and can lead to inadequate water provision and solid waste disposal which favour the proliferation of both vectors and virus transmission [13]. Increasing trade has facilitated expansion of *Aedes* vectors while increasing travel of humans has spread new viruses and virus sub-types into previously naive populations [14]. Finally, the level of local immunity also helps determine arboviral transmission patterns. Immunity is driven by both demography and past pathogen circulation patterns and can vary substantially between populations. The inherent spatial and temporal patterns of arbovirus transmission are therefore the result of the complex interactions of multiple factors, likely differing between arbovirus, location and spatial scale.

A wide range of spatial modelling techniques has been developed to account for complexities in investigating the variations in geographic spread of *Aedes*-borne arbovirus infections. Broadly, these can be categorised into i) data-driven approaches where flexible statistical models aim to recreate observed patterns with fewer built-in mechanistic assumptions about how variables are related to risk or ii) process-driven approaches where assumptions about drivers and how they affect transmission are encoded in a mechanistic (mathematical) model, which is then fit to observational data. Due to data scarcity in many risk mapping applications, implementing statistical and mathematical models in Bayesian frameworks has become increasingly popular due to incorporating prior information and better representing uncertainty in their predictions.

Previous systematic reviews have been conducted to identify and characterise dengue transmission models focused on predicting trends over time (hindcasting with the goal of developing forecasting systems) as opposed to spatially explicit prediction (risk mapping) [15–17]. Some of these systematic reviews included risk mapping studies but they have been limited to just a single arbovirus, usually dengue [7,18–20]. Although arbovirus risk mapping studies have become more diverse and advanced, to our knowledge, there are no systematic reviews that consider the important similarities and differences among arboviruses. Therefore, this study aims to identify and review studies that map *Aedes* mosquito-transmitted arbovirus risk in humans, and to characterise epidemiological data, covariates, modelling frameworks and methods of evaluation used.

## Methods

This review employed a search strategy and inclusion and exclusion criteria based on the preferred reporting items for systematic reviews and meta-analyses (PRISMA) guidelines [21].

### Search strategy

Four online bibliographic databases were searched: Embase, Global Health, Medline, and Web of Science. The final search was conducted on 15 June 2022 using institutional access from Oxford University. The search strategy included keywords and Medical Subject Headings (MesH) related to different arboviral diseases (namely dengue, Zika, chikungunya, and yellow fever) and those related to prediction. Search terms included “(Dengue OR DENV OR Zika OR ZIKV OR Chikungunya OR CHIKV OR Yellow fever OR YFV) AND (predict* OR forecast* OR map* OR driver*)”. Additionally, we manually searched the reference lists of articles and contacted experts in the field of arbovirus modelling to identify any studies not identified through the database search.

### Selection process

Results from database searches were combined and stored using Zotero referencing software; duplicates were removed using R (version 4.2.2) [22] by comparing the Digital Object Identifier (DOI) numbers of each study. Titles and abstracts were screened independently by two team members. All identified papers were included in full-text review and irrelevant articles were excluded. Full-text review was completed and disagreements on inclusion were resolved by consensus.

### Inclusion/exclusion criteria

Articles must be peer-reviewed, published in English and contain a spatial model that investigates the transmission of the arboviruses to humans. Spatial models were defined as models that included geographically realistic and explicit representations of more than one spatial location. While our primary focus was to review spatial models, spatiotemporal models were also included. There were no geographical or publishing date restrictions applied. We only included models that made predictions of some measure of the population-level virus infection risk, including but not limited to occurrence, incidence, prevalence, and proxies of transmission risk (e.g. reproduction number). Studies where the model was developed and/or validated in a previous paper were also included.

Articles were excluded if they only modelled transmission to vectors or non-human hosts or were exclusively dealing with occurrence of or suitability for the mosquito (e.g. vector suitability). Studies were excluded if they had only descriptive mapping of incidence using geographic information systems or if the model was not fitted or validated using observation data. Simulation-based and theoretical modelling studies were excluded unless their predictions of *Aedes-*borne disease transmission risk (as opposed to model parameters) were validated using data from real-world settings. Conference and workshop proceedings were excluded, as were review articles. This systematic review is registered on PROSPERO (reference: CRD42022358144).

### Data extraction

The following variables were extracted from eligible articles:

– study identification (title, author names, year of publication, study area, disease studied);
– model characteristics (type of model used, covariates included, covariates tested and not included, spatiotemporal resolution, assessment of collinearity);
– model validation (validation methods, metrics used to assess the model performance)

Analysis of the data and visualisations were carried out using R (version 4.2.2) [22]. The complete list of all included studies and data extracted from each study are available in S1 File.

### Quality assessment

A quality assessment tool was developed using the EPIFORGE checklist (S2 File), a guideline for standardised reporting of epidemic forecasting and prediction research, to assess the reporting quality of included studies [23]. This guideline assesses whether studies report on the following domains: study goals, data sources, model characteristics and assumptions, model evaluation, and study generalisability. The nine criteria were equally weighted, each with a score of 0 (poor) to 2 (good), for a maximum of 18 points. On the basis of the overall score, each paper was rated ‘low’ (<10), ‘medium’ (10–12), ‘high’ (13–15) or ‘very high’ (>15).

## Results

A total of 16,625 records were retrieved from the databases and 7,742 titles and abstracts screened after removing duplicates (Fig 1). A total of 83 records were additionally identified through bibliographic searches and contacts with experts. Of 301 records, a total of 118 studies were excluded because the full-text was not available, they were published in other languages, or the topics were irrelevant. One paper included two different models using different datasets so we counted it as two separate studies [24]. As a result, we identified 183 studies published between 2002 and 2022 that were ultimately included in the review (Fig 1).

**Fig 1.**
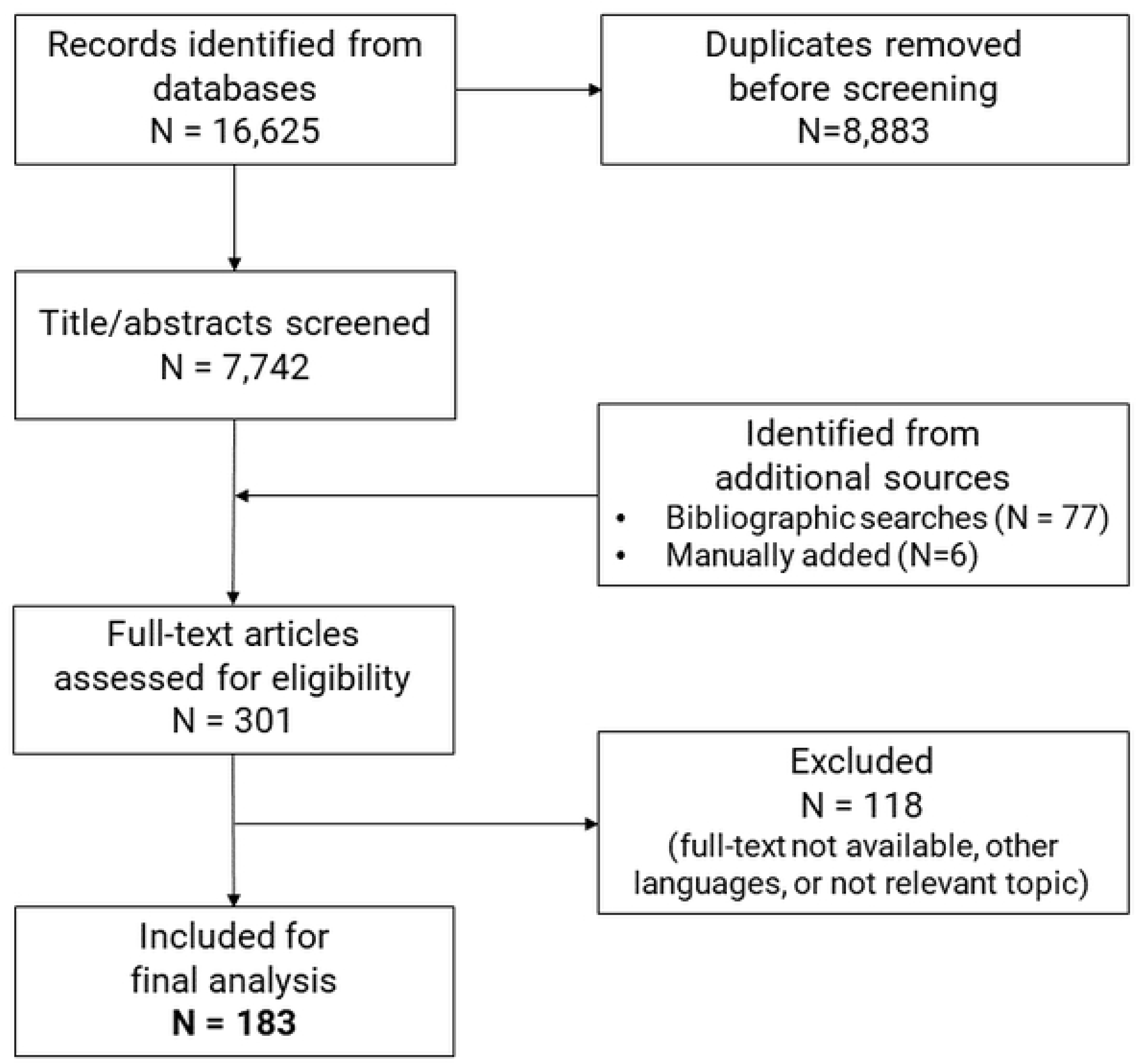
PRISMA flow chart.

There has been a rapid increase in the number of arboviral spatial modelling studies over the past 20 years, reflecting the growing public health priority of these diseases and increasing accessibility of data and modelling methods. There was an average of 1.7 studies published per year before 2008, 4.7 studies per year between 2008-2014 and 19.3 per year between 2015-2021 (Fig 2). The distribution of risk mapping studies over geography and by disease closely follow the abundance and availability of data. Using WHO Regions, a total of 40.8% (n = 78) of the studies were conducted in the Americas, followed by 19.4% (n = 37) in SE Asia and 17.3% (n = 33) in the Western Pacific region with a wide geographic diversity of studies over the past five years. Brazil (n = 35) was the most frequently studied country, followed by Colombia (n = 15) and Indonesia (n = 13). The diversity of regions studied has also increased: until 2014 studies tended to focus primarily on the Americas and Western Pacific whereas since 2015 studies focusing on SE Asia and the global scale have been increasingly prevalent (Fig 2). More than 70% (n = 131) of the studies modelled dengue transmission, 20 (10.9%) modelled Zika, 15 (8.2%) modelled yellow fever and seven (3.8%) chikungunya. There were six (3.3%) studies that modelled the risk of dengue, Zika, and chikungunya together, while also modelling the diseases individually; two modelled dengue and Zika together and two modelled Zika and chikungunya together.

**Fig 2.**
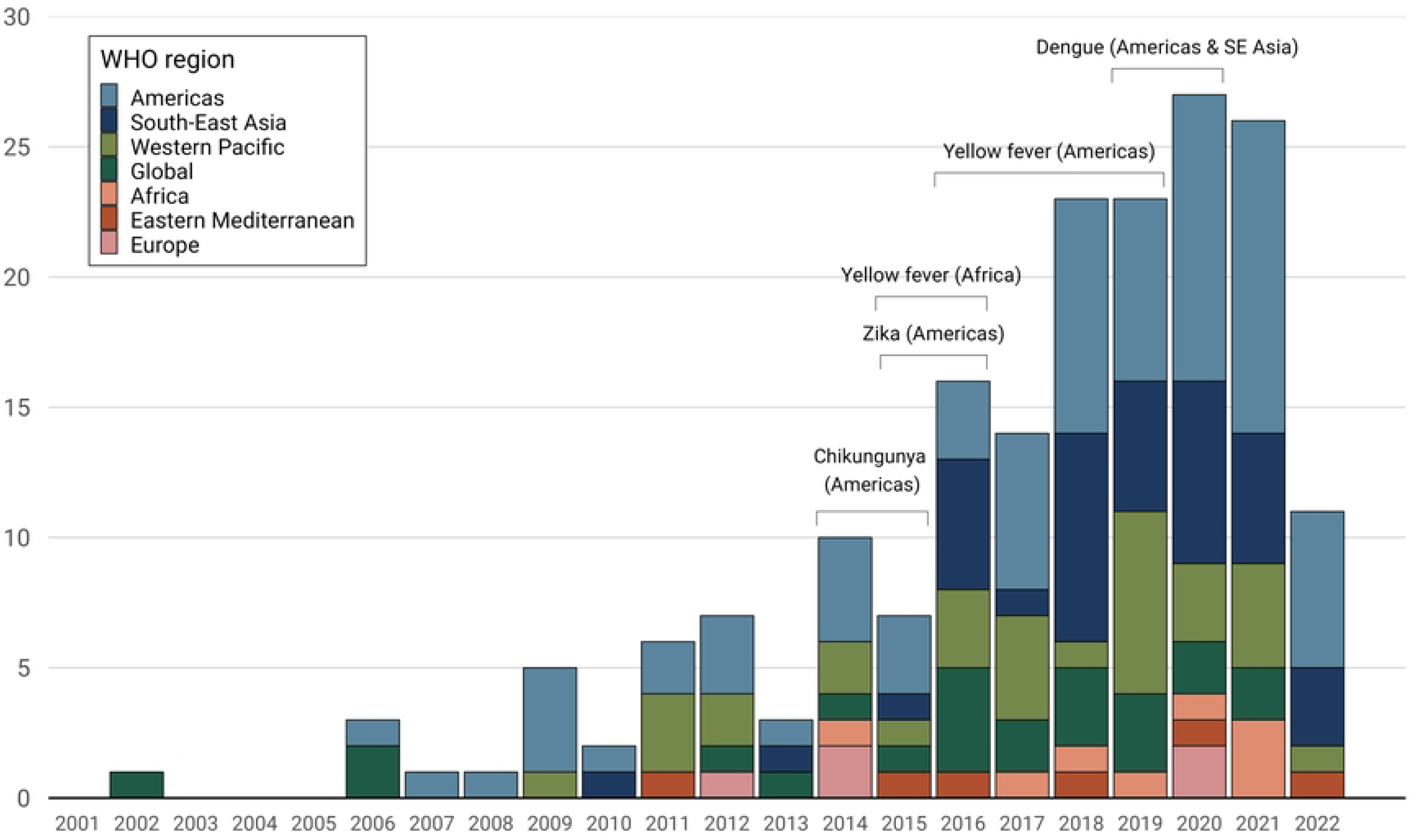
Number of included studies per year by study region. The brackets represent the key years for *Aedes*-borne arbovirus outbreaks, including chikungunya in the Americas (2014-2015) [25], Zika in the Americas (2015-2016) [7], yellow fever in Brazil (2016-2019) and Angola and Democratic Republic of Congo (2015-2016) [26], and dengue in the Americas & SE Asia (2019-2020) [27].

### Purpose of maps

The main groups of purposes or goals of risk maps vary depending on the specific disease and context, but can generally be grouped into four categories: 1) providing a broad overview of the spatial distribution of risk over long-term averages and suggesting how it might change under different scenarios of global changes in climate, economics, and demographics (e.g., [28,29]); 2) predicting the spread of outbreaks and gaining a better understanding of major drivers of geographical spread (e.g, [30,31]); 3) evaluating and planning vaccination programs by estimating disease burden and identifying high-risk areas at the continental or country-level scale (e.g., [32,33]); and 4) informing planning and outbreak response by increasing the precision of risk estimates and mapping sub-national risk using surveillance data (e.g., [34,35]).

### Data types

Most studies (n = 137, 74.9%) used case count data from routine passive surveillance to fit models, most often aggregated to the administrative district (admin2)-or province (admin1)-level (Fig 3). Use of occurrence data was also widespread (n = 29, 15.8%), particularly for specific use cases, such as the generation of global suitability maps. There were only seven studies (3.8%) that included data from community-representative seroprevalence surveys, and seven studies that included data from at least two different data types. The use of seroprevalence data was limited to dengue (n = 9) and yellow fever (n = 4), both resulting from widespread seroprevalence surveys in preparation for, or to evaluate, vaccination programmes. Generally the paucity of any one data type for yellow fever meant a more equally distributed use of different data types in models and greater use of multiple types of data [8,33,36,37] (Fig 3).

**Fig 3.**
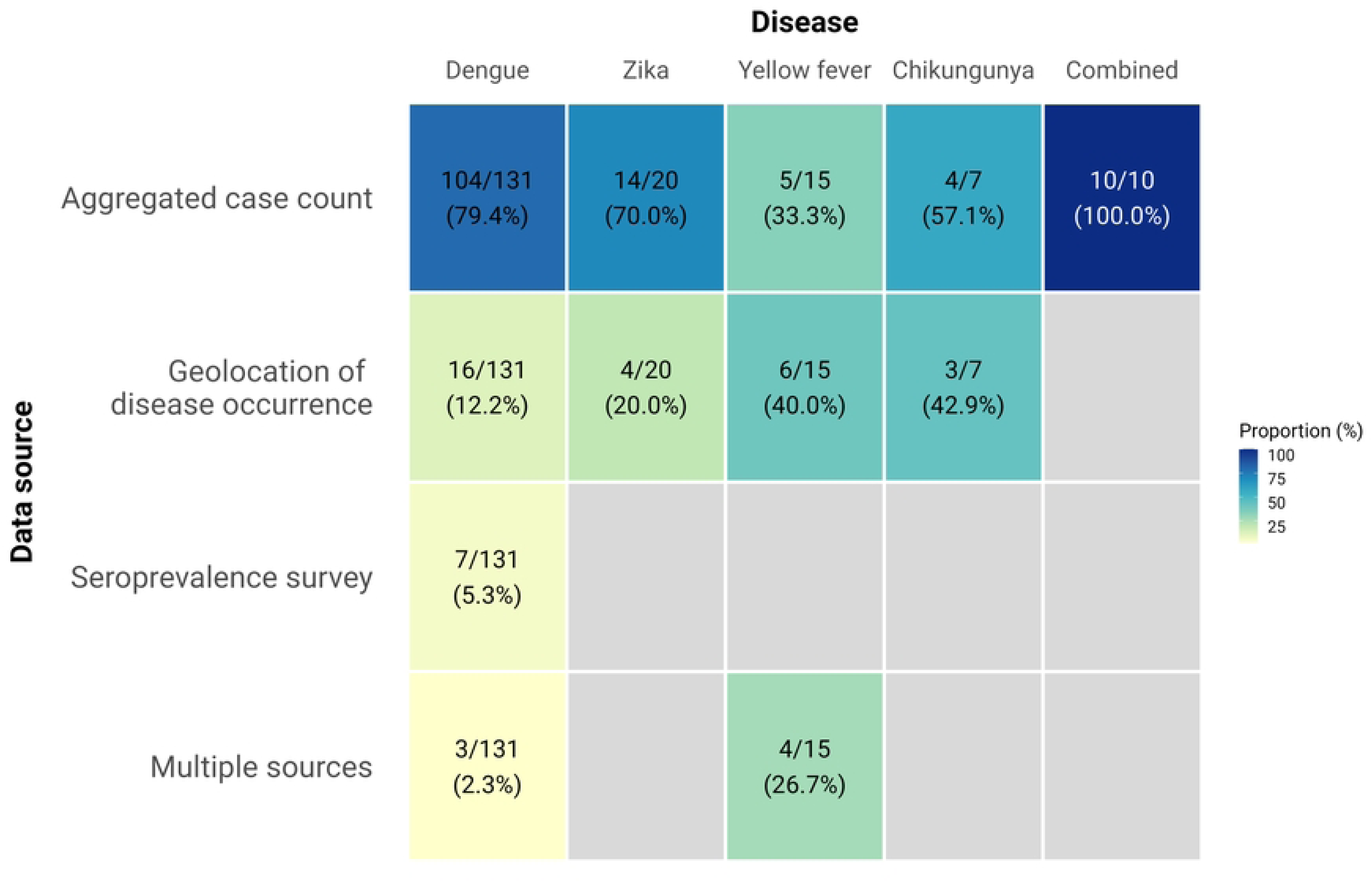
Sources of epidemiological data used by diseases. Each cell represents the number and percentage of studies with the denominators summed vertically.

Risk maps have been generated across a wide range of spatial scales from global to sub-national (Fig 4). We identified 22 studies that produced global risk maps of various *Aedes-*borne arboviruses. Despite large gaps in data availability at the global scale, the majority (n = 18/23, 78.3%) of these global maps make high resolution predictions at the pixel level, enabled by growing availability of high resolution remotely-sensed climate datasets (Fig 5). For Zika, yellow fever, and chikungunya, maps were primarily focussed at a continent or national scale with a resolution between city-level and national-level (Figs 4 and 5), reflecting the more regional scope of their distribution (yellow fever in Africa) or high profile epidemics (the 2015-2016 Zika epidemic in the Americas). While maps are available at all spatial scales for dengue, the majority of models (n = 83, 63.4%) are now at sub-national scale, usually at the resolution of city/district (admin-2) (Figs 4 and 5). This reflects the increasing application of these techniques to routinely collected case incidence data to provide country-specific recommendations about targeting of control resources within countries based on the latest local data. There remain strong regional disparities in the scale and resolution of mapping efforts with many high-resolution and country-specific maps in the Americas, while risk estimates for Africa are fewer, of comparatively lower resolution, and are typically derived from global or continent-level modelling efforts (S1 Fig).

**Fig 4.**
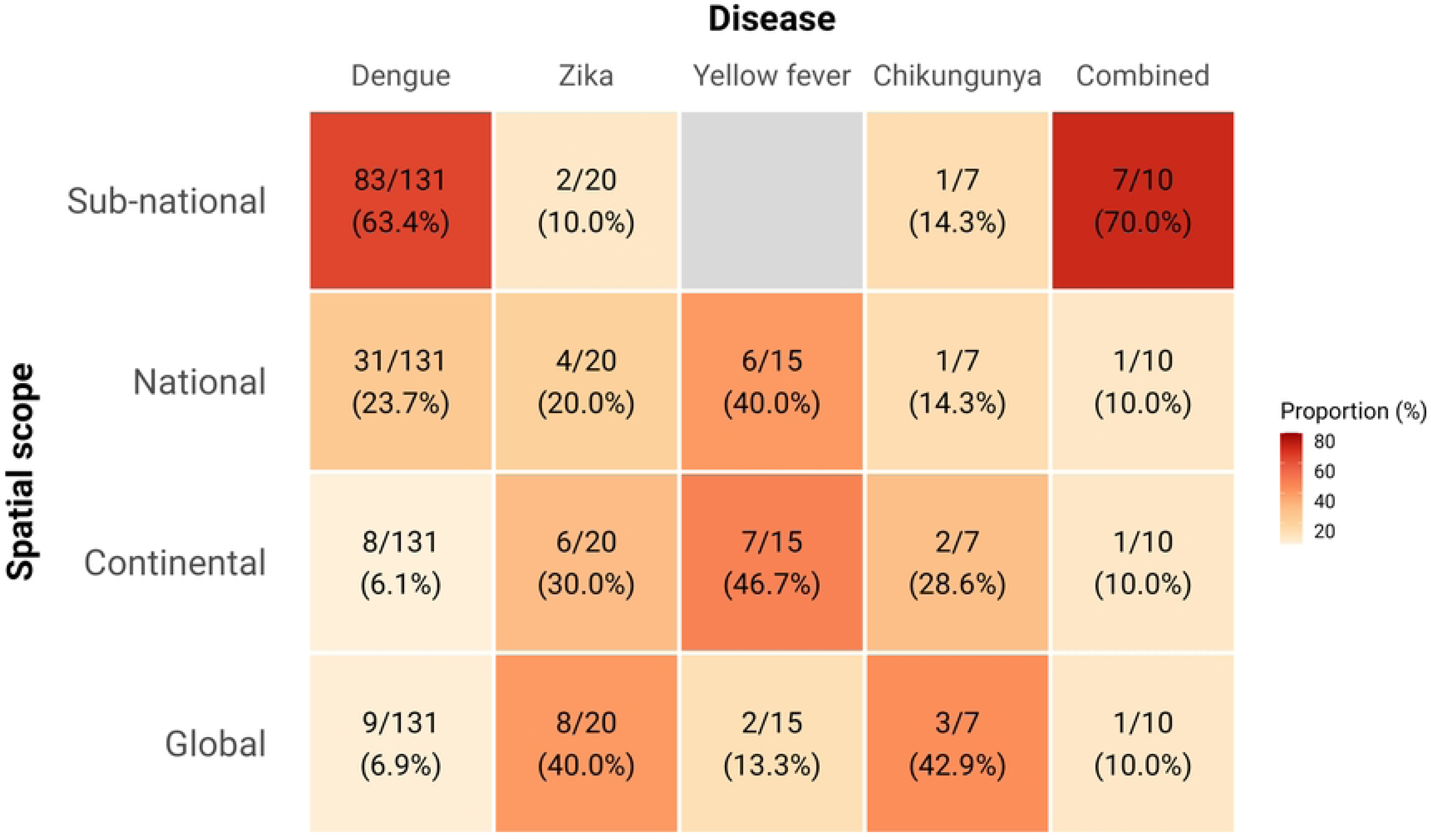
Geographical scope by diseases. Each cell represents the number and percentage of studies with the denominators summed vertically.

**Fig 5.**
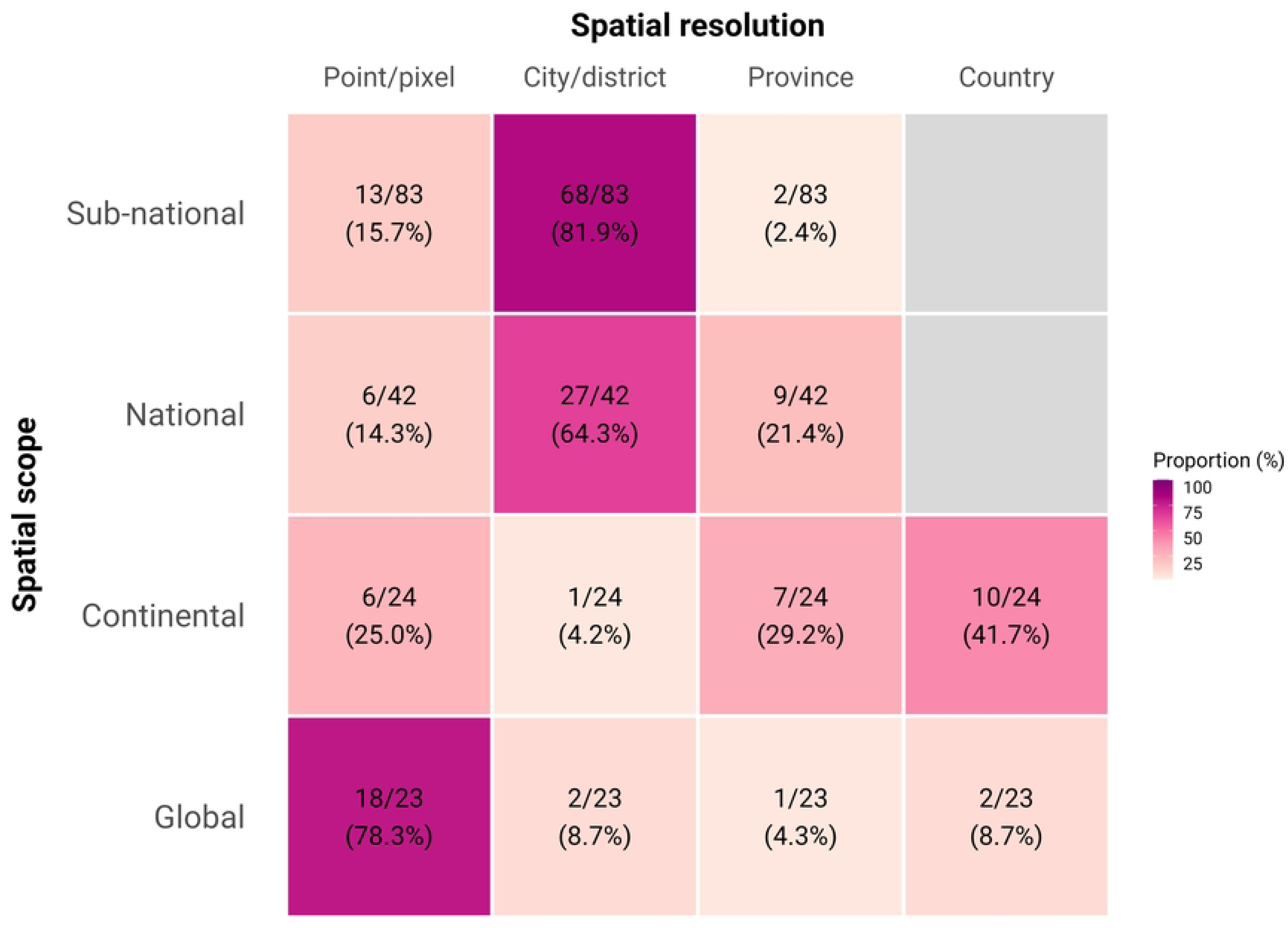
Spatial resolution by geographical scope. Each cell represents the number and percentage of studies with the denominators summed horizontally.

Spatiotemporal prediction maps were often generated based on monthly or weekly intervals (S1 Table). The longest period of study was for 804 months (67 years), while the shortest period of study was for 3 months, with an average of 125 months (10 years) and a median of 60 months (5 years). Studies tended to use data from periods with high numbers of reported cases, with dengue data concentrated in between 2010-2015, Zika data between 2015-2016. For chikungunya many studies use data from 2014 and for yellow fever the data used have been spread over time, with few studies using recent data from 2015-2020 (S2 Fig).

### Covariates

Studies reviewed included a wide range of covariates in their models (Table 1). We grouped these into six main groups: climatic, demographic, socioeconomic, ecological, environmental and spatiotemporal incidence.

**Table 1.** List of covariates included in the studies.

| Covariates | Count | Percentage (%) |
| --- | --- | --- |
| **Climatic** |  |  |
| Temperature | 98 | 53.6 |
| Rainfall | 79 | 43.2 |
| Humidity | 23 | 12.6 |
| Bioclimatic variables | 6 | 3.3 |
| El Nino Southern Oscillation Index | 4 | 2.2 |
| Soil moisture (water stress/wetness) | 4 | 2.2 |
| **Demographic factors** |  |  |
| Population density | 44 | 24.0 |
| Age | 26 | 14.2 |
| Air travel | 19 | 10.4 |
| Human daily mobility | 13 | 7.1 |
| Vaccination coverage | 7 | 3.8 |
| Sex | 7 | 3.8 |
| **Socio-economic factors** |  |  |
| Gross domestic product | 15 | 8.2 |
| Household income | 12 | 6.6 |
| Education/literacy rate | 6 | 3.3 |

| **Covariates** | **Count** | **Percentage (%)** |
| --- | --- | --- |
| Occupation and employment status | 5 | 2.7 |
| Socio-economic strata | 6 | 3.3 |
| **Ecology** |  |  |
| Non-human primates species | 6 | 3.3 |
| Location of breeding sites | 6 | 3.3 |
| Breteau index | 3 | 2.2 |
| Adult mosquito abundance | 2 | 1.1 |
| **Environmental factors** |  |  |
| Vegetation | 27 | 14.8 |
| Elevation/altitude | 25 | 13.7 |
| Urbanisation | 22 | 12.0 |
| Distance to roads, road density | 14 | 7.7 |
| Land use/land cover | 13 | 7.1 |
| Distance to water bodies/river | 9 | 4.9 |
| **Spatiotemporal incidence** |  |  |
| Case count across time periods and neighbouring regions | 23 | 12.6 |

Climatic variables were the most common group of covariates in models with temperature and rainfall dominating. More than half of the studies (n = 98, 53.6%) included temperature as a covariate while around 40% of studies had rainfall (n = 79, 43.2%). Temperature and rainfall were better fit when lagged one or two months rather than unlagged [38–41]. Temperature and rainfall were considered as significant factors in most studies, but some studies showed that meteorological factors alone are not sufficient to explain spatial heterogeneity in disease transmission, which may be associated more with non-climatic factors [42–44]. Rather than rely on raw measures of temperature, 24 studies (13.1%) instead used “temperature suitability” of *Aede*s mosquito vectors, which incorporates a variety of different methods of modelling the temperature constraints on the vector and virus dynamics that are most critical for virus transmission [45]. Six studies used bioclimatic variables that encompassed annual temperature and precipitation ranges, seasonal fluctuations, as well as extreme or constraining factors that capture broader biological patterns [29,46–50]. Four studies additionally used indicators associated with El Niño Southern Oscillation as covariates [35,51–53]. Examples of other climatic variables that were included in the reviewed models were diurnal temperature range [54–56], atmospheric pressure [57,58], wind speed [59,60], and duration of sunshine [38,61,62].

Population density (n = 44, 24.0%) and age distributions (n = 26, 14.2%) were often considered in modelling arboviruses. Many studies found population density to be a significant covariate in their models, demonstrating a positive association with disease transmission, but some studies reported a negative [63,64] or null association [39,44,65]. Human mobility between cities or countries (n = 19, 10.4%) was also considered by including travel distance between regions [66,67] or air travel passenger volume [68–73]. Some studies included daily human mobility data (n = 13, 7.1%), mostly mapped at sub-national scale, with the aim of better representing short-distance high frequency movements such as daily commuting [74,75]. Seven studies, for yellow fever and dengue, considered vaccination coverage and measures of population immunity from infection in their models [27,33,37,47,63,76,77].

The most common socio-economic variable was gross domestic product (GDP) (n = 15), followed by household poverty/income level (n = 12, 6.6%) and education level (n = 6, 3.3%). A socio-economic strata or a composite index such as human development index, social advantage and disadvantage score (n = 6, 3.3%) were also included as socio-economic predictors in some of the reviewed models. Lower neighbourhood socio-economic status was generally associated with increased risk of *Aedes-*borne arbovirus diseases; in regions with established arboviral circulation, community-level factors such as inadequate garbage collection, low income, and lack of access to health care were associated with elevated risk of dengue infections [78–80].

For models fit at the sub-national scale to case incidence data, accompanying direct measurements of the *Aedes* mosquito population improved model predictive performance. Breteau index (BI), which is defined as the number of positive containers per 100 houses, was used as a predictor in three studies [53,81,82]. Six studies included location of *Aedes* breeding sites in their models [74,83–87]. The number of catches of female adult mosquitoes was included in two studies [58,88]. In the absence of direct measurements of the vector abundance, modelled predictions of “suitability for *Aedes* mosquitoes [89]” have been used, particularly at broad global scales and to make early predictions for emerging Zika epidemics. Six studies included the occurrence or species richness of non-human primates in modelling yellow fever.

The most common environmental variable was vegetation index (n = 27, 14.8%), followed by altitude/elevation (n = 25, 13.7%) and urbanisation (n = 22, 12.0%). Some studies found that vegetation was not a key predictor variable and had no association with dengue incidence [90,91], whereas those considering vegetation in modelling yellow fever generally found that there was a strong and significant vegetation-disease association possibly because of the greater role of the forest-fringe environment in driving spillover from non-human primate reservoirs [64,92–94]. Road density and proximity to the road were also included as a predictor in 14 studies (7.7%). More generic categories of land use and land cover type have also been considered in another 13 studies.

Disease incidence across time periods and neighbouring regions were included as covariates in 23 studies (12.6%) to explain contemporaneous disease transmission. Several studies included past case counts lagged by one week to four months to improve temporal prediction accuracy [51,66,95–98]. Source country’s disease incidence rate was included in studies quantifying the risk of importation from endemic to non-endemic settings such as Europe [70,99] and Asia-Pacific regions [69].

For each paper, we also examined whether the collinearity among covariates was checked and whether models retained covariates after conducting variable selection procedures. Among the 148 studies excluding those that used mechanistic models or only included random effect terms, only 33 studies (22.3%) tested different combinations of covariates and checked the multicollinearity among them by calculating the correlation coefficient or variance inflation factor, or using principal component analysis. There were 63 studies (42.6%) that did not include any process for selecting variables or checking collinearity (S3 Table). However, it is worth noting that some of these studies may have had a small number of covariates that were selected based on their known or cited ecological or theoretical relevance to disease transmission, which may explain the lack of variable selection process.

For the 33 studies that both checked the multicollinearity of covariates and performed variable selection, we summarised the retention rate of different groups of covariates in the final models (Fig 6A) [27,32,38–44,46–48,54,59,60,72,78,79,86,93,99–112]. Of 33 studies, 25 studies (96.2%) retained climatic variables when tested. Only one study on dengue [111] tested all six categories and rejected demographic, ecological data and spatiotemporal incidence; seven studies tested all categories except for ecological variables. Apart from climatic variables, environmental variables were the most commonly used, with 21 studies tested and only three of them rejected, followed by demographic (23 tested and 6 rejected), socio-economic variables (16 tested but 5 rejected). Ecological data (7 tested and 2 rejected) and spatiotemporal incidence (5 tested and 1 rejected) were the least tested and included (Fig 6A). The most common combinations of retained categories were climatic, environmental, demography, and socio-economic (n = 4) [41,44,103,108]. For climatic variables, different measures of temperature and rainfall were tested in reviewed studies. Inclusion of temperature in models differed between studies, with minimum temperature often selected over average and maximum temperature in six out of 14 studies (Fig 6B). We identified that average rainfall was preferred over other measures of rainfall and humidity but only five studies examined the performance of models in which both variables were considered (Fig 6B). We found that 29 studies have included lagged covariates in their models. The length of the lag periods tested for temperature, humidity and precipitation ranged from 0 to 16 weeks, with most being concentrated between 4 to 12 weeks (S3 Fig). The average lag periods for mean temperature and precipitation tend to be longer in the Americas compared to Western Pacific and SE Asia (S3 Fig).

**Fig 6.**
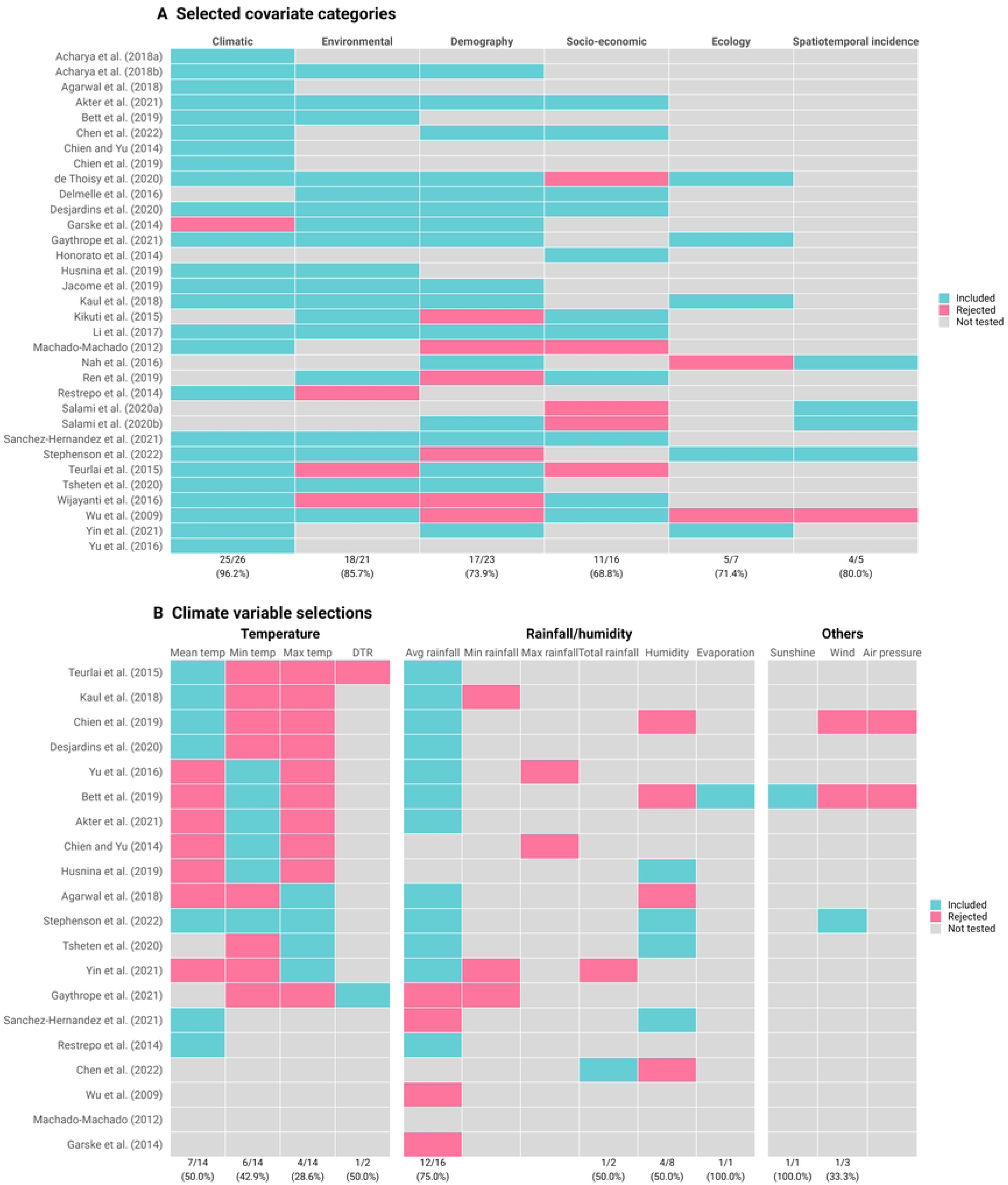
Covariates included and rejected. (a) Selected covariate categories; (b) climate variable selections. Mean temp: mean temperature; Min temp: minimum temperature; Max temp: maximum temperature; DTR: Diurnal temperature range; Avg: average. The values in the bottom represent the number and percentage of studies tested and included the corresponding category of covariates.

### Modelling framework

Four classes of modelling methods were identified: statistical mixed effect models, statistical fixed effect models, machine learning and mechanistic models (Table 2). Overall, the most common modelling approaches were types of statistical mixed effect models (n = 69, 39.5%), with generalised linear mixed models (GLMM) dominating, followed by generalised additive mixed models (GAMM) (n = 4) and distributed lag non-linear models (DLNM) (n = 4). Mixed effect models were often preferred when using areal-type case count data aggregated over distinct geographical areas (e.g. administrative boundaries) (Fig 7).

**Fig 7.**
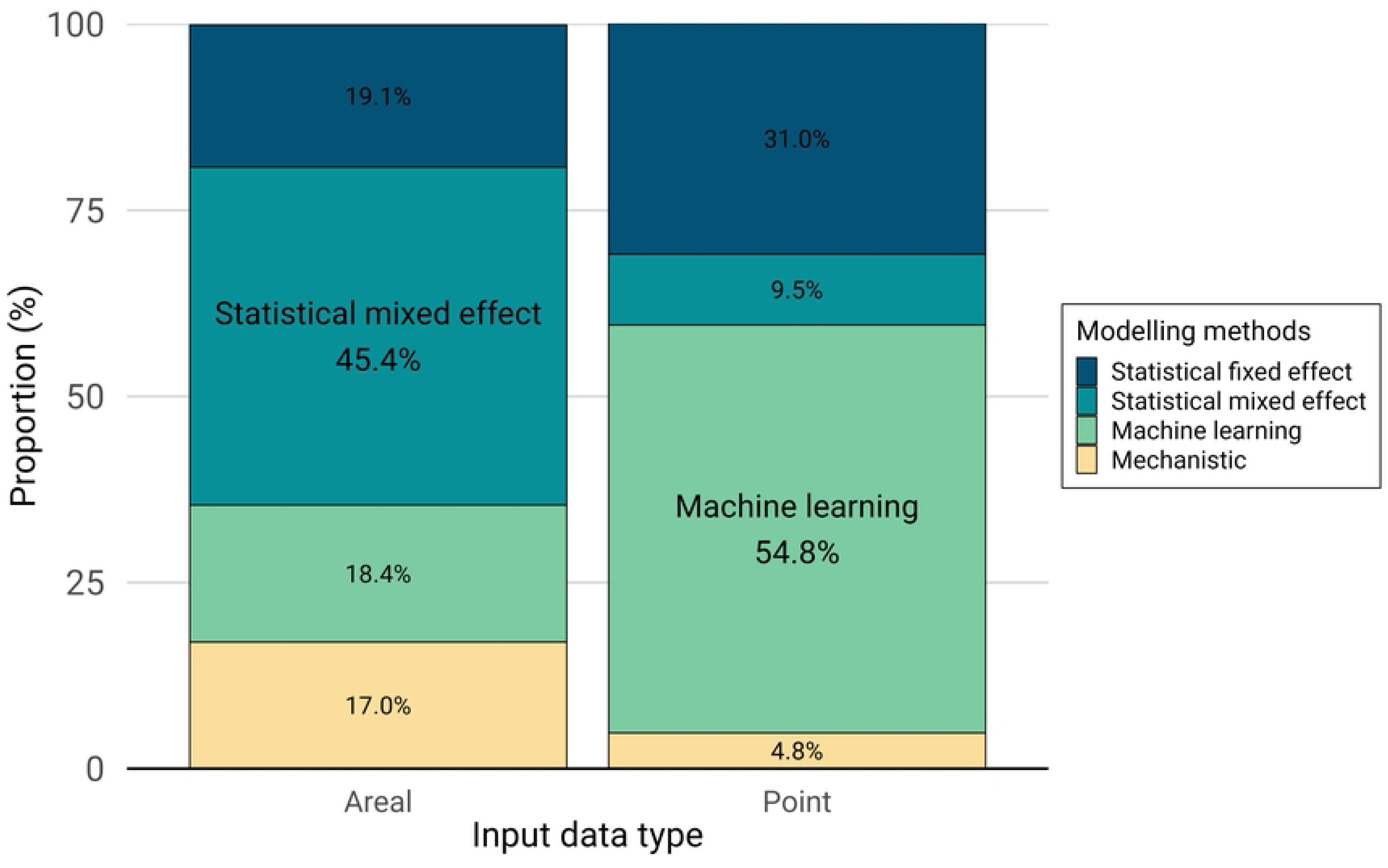
Modelling framework by input data type.

Statistical fixed effect models were used in 21.2% of studies, with generalised linear models (GLM) and geographically weighted regression (GWR) as the most used approaches. Since fixed effect models assume that all observations are independent, models used spatial variables to account for spatial relationships. For example, several studies included the coordinates (long, lat) of cases, households, or the centroid of a region [32,36,77,113–116].

A variety of machine learning methods were employed in 26.1% of studies. The most frequently used machine learning methods were MaxEnt and boosted regression tree (BRT). They were often used when developing ecological niche or species distribution models using point-referenced occurrence data to describe the environmental suitability of arbovirus transmission, and especially for larger geographical scales (e.g., international scale). Of 23 studies that developed a global risk map of different arbovirus transmission, ten studies adopted machine learning methods, six of which used either MaxEnt or BRT [3,28,29,117–119]. Seven studies developed and compared the performance of different machine learning methods. For example, Jiang et al. (2018) adapted three different machine learning models, namely backward propagation neural network, gradient boosting machine and random forest, and reported that backward propagation neural network showed the best performance in predicting the global transmission risk of Zika [120]. Two studies generated ensemble model projections of the spatiotemporal dynamics of Zika in Brazil and burden of yellow fever in Africa [121].

Mechanistic models were used in 15.2% of studies, especially compartmental and metapopulation models. Compartmental models e.g. human SEIR – mosquito SIR models were used in six studies to explain the impact of different factors on the transmission dynamics, especially for smaller scales e.g. country or sub-national scale [75,88,122–125]. Eight studies used metapopulation or network models, all of which considered the connectivity between areas or regions by including the patterns of daily human mobility or air travel data [31,67,70,71,73,82,83,126]. Five studies used mechanistic mosquito models to produce estimates of temperature suitability, vectorial capacity or basic reproductive number (R0) at the continent or global scale [127–131].

Surprisingly, only 48.1% of studies (n = 88) included in this review assessed the predictive performance using cross-validation procedures, such as K-fold cross-validation or random partitioning of data, commonly referred to as “out-of-sample validation”. It was more common to perform this type of validation in studies using machine learning methods than in studies using other modelling methods; only 25% of studies using fixed effect models performed out-of-sample validation (Fig 8). Of these studies, only three studies included model validation on independent test data (“hold-out validation”) [55,132,133].

**Fig 8.**
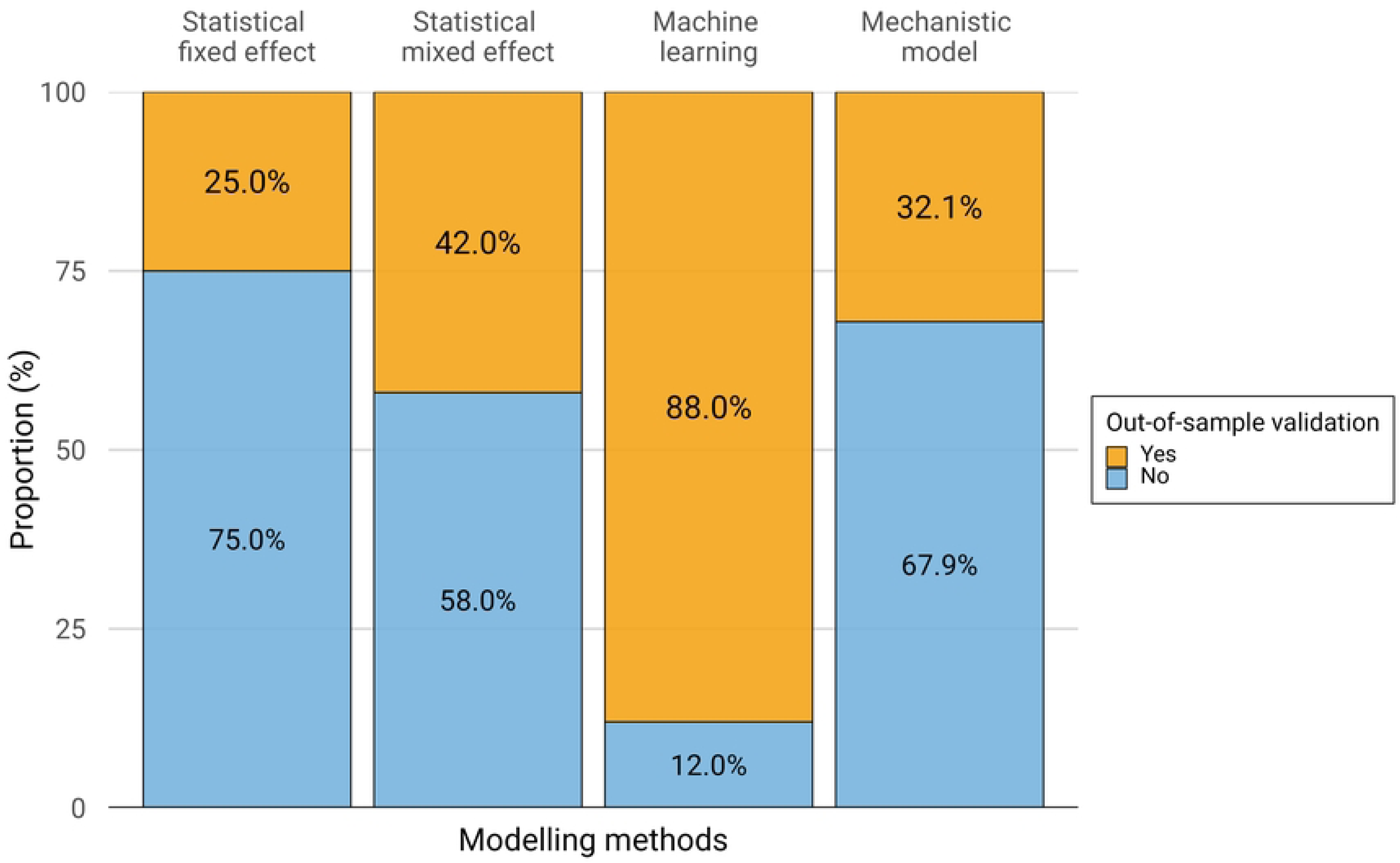
Out-of-sample validation by modelling framework.

The most common model performance evaluation metrics were information criteria (n = 82, 29.8%), with Akaike information criteria (AIC) and the Bayesian information criterion (BIC) as the most used metric (S2 Table). Confusion matrix-based metrics were used in 21.1% of studies, with the Receiver Operating Characteristics (ROC) curve most frequently used. Correlation-based metrics were used in 14.9% of studies, especially R-squared. 23 studies (8.4%) did not use any of the metrics described above (S2 Table).

### Quality assessment

Using the adapted tool for assessment of modelling study reporting, scores for the reviewed paper ranged from 6 to 18 out of 18. Twelve studies were classified as low quality, 50 as medium quality, 76 as high quality and 45 as very high quality. The median score was 13/18, which is categorised as high quality. Discussions on the generalisability of the developed models were lacking in many papers. Study objectives, settings, and data sources were often unclear in poorly scored studies.

## Discussion

This review provides a comprehensive overview of risk mapping studies, including their covariates and modelling frameworks to investigate the transmission of arboviruses. We found that the choices of data, covariates and modelling frameworks were largely determined by the purpose of the map. We identified 23 studies that generated global risk maps, using machine learning-based ecological niche modelling. These approaches are designed to give a broad overview of the spatial distribution of risk over long-term averages and suggest how it might change under different scenarios of global changes in climate, economics and demographics. Geolocation of disease occurrence data, often combined with high-resolution environmental datasets, were more common for global risk mapping because they encompass large areas and various environmental conditions and provide information about the extent of transmission. However, caution is needed when utilising the outputs of high resolution global risk maps, particularly for informing local decisions due to large data gaps and biases [133] that are not reflected in their highly geographically precise predictions and sometimes don’t align with (typically later published) estimates from country-specific models that use more local data.

We found that major epidemics, such as the 2015-2016 Zika epidemic, have acted as catalysts for the development of new risk mapping methods applied in new contexts, possibly due to expanding generation and sharing of data that has accompanied these more recent epidemics. The paucity of data in the early stages of epidemics and similarities between arboviral diseases gives mechanistic modelling approaches an advantage over more data-dependent statistical approaches despite the latter’s traditional dominance of the field of risk mapping [14]. As with any model, the predictions are inherently a function of the data available and primary use cases at the time of analysis, and contemporary approaches to mapping risk of diseases like Zika and chikungunya would likely differ substantially from those conducted in the early stages of epidemics. We also show how epidemics have accelerated the use of human movement data in arbovirus risk mapping, and that human movement data is especially valuable to understand long-distance spread since *Aede*s mosquitoes have a limited dispersal capability [134]. Daily commuting and air travel has improved predictions in both statistical and mechanistic modelling approaches, particularly when mapping how the spatial distribution of risk changes over the course of an epidemic.

Studies on modelling yellow fever employed multiple datasets and various approaches, mostly motivated by the need to account for sparse, non-standardised data. They tend to be conducted at continental or country-level scale in African and South American countries with high endemicity for yellow fever transmission or recent outbreaks, for the purpose of evaluation and planning vaccination programs. Inclusion of seroprevalence data and vaccination coverage therefore played a significant role in robust estimation of disease burden and approaches used for yellow fever could be increasingly important for mapping dengue risk as vaccines begin to be rolled out in various countries [135].

In contrast, the majority of publications that use predictive risk mapping for dengue (which accounted for more than 70% of the studies included in this review) now focus on mapping sub-national risk using case incidence data from a country’s passive surveillance system. Such models theoretically offer the most potential for direct integration with country surveillance systems and would allow risk maps to directly inform planning, intervention targeting and outbreak response. The proliferation of risk mapping in this domain closely aligns with improvements in routine dengue disease surveillance and sharing of sub-nationally disaggregated data and could be applied to other emerging disease threats if similar approaches to surveillance are adopted. We found that statistical mixed effect models were more commonly implemented than machine-learning approaches for sub-national models, which allow more constraints over the effects of environmental covariates and are easier to implement in Bayesian frameworks, both assets that allow more stability and better representation of uncertainty when making spatio-temporal predictions. Such models blur the boundaries between pure risk mapping (predicting to new spatial locations) and pure hindcasting/forecasting (predicting to new periods of time) and show the added value considering both spatial and temporal information can contribute to each of these applications.

Overall, we found that the quantity and variety of covariates included in arbovirus risk mapping studies has increased in line with growing availability of these variables. While developments over the past decade have focussed on global climate datasets, data on human movement [136] and urban infrastructure [137] are becoming increasingly available and may play important roles in future arbovirus risk mapping studies. Historically, limited data availability has made it difficult to quantify human mobility patterns, requiring models that incorporate gravity or radiation as an approximation [31,83,138]. However, the recent emergence of mobile phone data enables real-time tracing of fine-scale movement across large numbers of individuals, although privacy and bias issues remain [139]. The move towards large, open, accessible datasets for vector borne diseases necessitates not just a more robust data science workforce, but a better motivation and capacity planning for data fluency among primary data producers. While issues of human subjects and data privacy must remain foremost in contemplating large-scale studies of vector borne disease risk, nonetheless, leveraging entomological surveillance data, meteorological data, geospatial representation of infrastructure and landscape (e.g., derived from remote sensing, well-resolved built environment enumerations, high resolution travel network data), and climatological modelling output, is less constrained by international regulations, so identifying the necessary investments and key routes of engagement is a high-level first step to addressing the data gaps.

We found surprisingly few studies conducted robust variable selection procedures. In addition, out-of-sample validation techniques were explicitly stated in only half of the studies reviewed. Statistical and machine learning models, predominantly used in arbovirus risk mapping studies, require a large amount of data and therefore both variable selection and cross-validation are important steps to reduce overfitting and improve model interpretability and predictive accuracy. Although the majority of studies used traditional cross-validation techniques, the use of spatial cross-validation i.e., spatial block bootstrapping is increasingly popular due to its ability to account for spatial dependence in the data [92,94]. This may help to better test the spatial predictive performance of the model, particularly if there are large heterogeneities in data availability across the study sites, which is common in many arbovirus mapping applications.

### Limitations

One limitation of our systematic review is that it focussed on spatial modelling approaches. The conclusions we reach, particularly with reference to drivers of transmission, may differ between risk mapping and temporal prediction models which may be particularly important as the two fields continue to overlap. We also only considered studies published in English, which may affect our conclusions about regional patterns. Additionally, it is possible that some relevant literature, particularly in the form of grey literature, may have been missed as the databases do not contain all journals and university press articles. This is particularly true for locally-relevant geospatial modelling work, which may not have been published in mainstream academic outlets. Finally, we excluded studies that did not assess risk of human infection, excluding a number of studies dealing exclusively with entomological risk or non-human host risk.

### Recommendations for future studies

● Consider the strengths and weaknesses of different data types for different purposes as the choice of data type imposes specific restrictions on the modelling framework and resolution of the prediction. Historically the most common applications have been: occurrence data to map the changing global limits of transmission, short-term aggregated level incidence data to track the geographic spread of epidemics and high spatiotemporal resolution incidence data to understand the roles of different drivers and forecast epidemics.
● Include covariates from multiple domains (climatic, environmental, demographic, socioeconomic, ecological) and test whether their inclusion improves prediction.
● National or subnational studies should consider additional local covariates not available across broader regions, such as data from the arbovirus control programmes, finer scale meteorological resolution data, or infrastructural data from census databases.
● Even with extensive use of covariates, unobserved confounding will still be an issue, particularly for broad scope (national and above) models, meaning that the use of structured spatio-temporal random effects, ideally in a Bayesian mixed effects statistical modelling framework, is preferable to more simplistic fixed effect statistical models.
● Use predictive validation metrics on held out datasets. Ideally using procedures that take into account the unique challenges posed by highly spatially and temporally heterogeneous datasets, such as multiple-fold blocked spatial and temporal cross validation.
● Arbovirus risk mapping is a rapidly developing field with continual improvements in data quantity and representativeness, growing availability of potentially informative covariates and new innovations to model fitting and evaluation. Future arbovirus risk mapping studies should incorporate these new developments and not just rely on the status quo of existing studies.

## Conclusion

Spatial modelling can help identify potential risk factors for arbovirus transmission and provide a better understanding of the current and future distribution of arboviruses. We provide a synthesis of covariates and modelling frameworks used for risk mapping of arbovirus, providing an evidence base for developing up-to-date arbovirus risk maps based on current best practices. Although approaches to map arbovirus risk have diversified, it is important to select the data, covariates, models, and evaluation methods based on the purpose of maps, data availability and epidemiological contexts.

## Data Availability

All relevant data are within the manuscript and its Supporting Information files.

## Acknowledgements

This work was discussed with the Technical Advisory Group on arboviruses (TAG-Arbovirus), the Secretariat of the Global Arbovirus Initiative (Raman Velayudhan, Laurence Cibrelus, Jennifer Horton, Marie-Eve Raguenaud, Maria Van Kerkhove, Qingxia Zhong), and the participants of the arbovirus risk mapping meeting held in Seattle in October 2022 as part of the ASTMH (Isabel Rodriguez-Barraquer, Leo Bastos, Simon Cauchemez, Ilaria Dorigatti, Neil Ferguson, Simon Hay, Wenbiao Hu, Axel Kroeger, Velma Lopez, A. Townsend Peterson, Maile Philips, David Pigott, Krystina Rysava, Sophie von Dobschütz, and Anna Winters).

## Supplementary information

S1 Fig. Spatial scale (a) and resolution (b) by study region. Each cell represents the number and percentage of studies with the denominators summed vertically.

S2 Fig. Time span of data used by disease.

S3 Fig. Summary of lagged covariates used. (a) lagged week per covariate; (b) average lag period of climatic covariates by region. The numbers represent the mean (standard deviation) of the lag period in weeks.

S1 Table. Temporal resolution of predictions in reviewed studies.

S2 Table. Modelling methods used in arbovirus risk mapping.

S3 Table. Number of studies that used robust variable selection procedures.

S1 File. Data extracted from the studies reviewed.

S2 File. A modified EPIFORGE checklist.

